# A highly effective reverse-transcription loop-mediated isothermal amplification (RT-LAMP) assay for the rapid detection of SARS-CoV-2 infection

**DOI:** 10.1101/2020.06.30.20142935

**Authors:** Veronica L. Fowler, Bryony Armson, Jose L. Gonzales, Emma L. Wise, Emma L. A. Howson, Zoe Vincent-Mistiaen, Sarah Fouch, Connor J. Maltby, Seden Grippon, Simon Munro, Lisa Jones, Tom Holmes, Claire Tillyer, Joanne Elwell, Amy Sowood, Oliver de Peyer, Sophie Dixon, Thomas Hatcher, Helen Patrick, Shailen Laxman, Charlotte Walsh, Michael Andreou, Nick Morant, Duncan Clark, Nathan Moore, Rebecca Houghton, Nicholas Cortes, Stephen P. Kidd

**Author notes:** Joint first authorship (Veronica L. Fowler, Bryony Armson).

## Abstract

The COVID-19 pandemic has illustrated the importance of simple, rapid and accurate diagnostic testing. This study describes the validation of a new rapid SARS-CoV-2 RT-LAMP assay for use on extracted RNA or directly from swab offering an alternative diagnostic pathway that does not rely on traditional reagents that are often in short supply during a pandemic.

Analytical specificity (ASp) of this new RT-LAMP assay was 100% and analytical sensitivity (ASe) was between 1×10^1^ and 1×10^2^ copies per reaction when using a synthetic DNA target. The overall diagnostic sensitivity (DSe) and specificity (DSp) of RNA RT-LAMP was 97% and 99% respectively, relative to the standard of care rRT-PCR. When a C_T_ cut-off of 33 was employed, above which increasingly evidence suggests there is a low risk of patients shedding infectious virus, the diagnostic sensitivity was 100%. The DSe and DSp of Direct RT-LAMP (that does not require RNA extraction) was 67% and 97%, respectively. When setting C_T_ cut-offs of ≤33 and ≤25, the DSe increased to 75% and 100%, respectively, time from swab-to-result, C_T_ < 25, was < 15 minutes.

We propose that RNA RT-LAMP could replace rRT-PCR where there is a need for increased sample throughput and Direct RT-LAMP as a near-patient screening tool to rapidly identify highly contagious individuals within emergency departments and a care homes during times of increased disease prevalence; ensuring negative results still get laboratory confirmation.

**Highlights:**

- Novel rapid RT-LAMP assay with diagnostic sensitivity and specificity of 97% and 99%
- Development of an RNA extraction free direct detection method for SARS-CoV-2
- Use case modelling for rapid Direct RT-LAMP in near-patient clinical practice
- Developing diversity of testing modalities within a diagnostic laboratory during a pandemic.

## 1. Introduction

In December 2019, an unusual cluster of pneumonia cases were reported by the Chinese Centre for Disease Control (China CDC) in the city of Wuhan, Hubei province^1^ It was quickly established by sequencing of airway epithelial cells that these patients were infected with a novel *betacoronavirus*^2^ which was named by the International Committee on Taxonomy of Viruses as severe acute respiratory syndrome coronavirus 2 (SARS-CoV-2) due to the close genetic relatedness to SARS-CoV^3^. Since its first discovery, SARS-CoV-2 has spread around the globe reaching pandemic status, and by June 2020 has infected 30.6 million people and caused more than 950 000 deaths according to The World Health Organisation situation report (accessed 24^th^ September 2020).

Genomic regions suitable for targeting with molecular tests such as real-time reverse-transcription polymerase chain reaction (rRT-PCR) were published by Corman *et al*^4^ early in the outbreak and comprised the *RdRp, E* and *N* genes. Diagnostic tests developed targeting these regions have since been utilised for routine use in many reference and hospital laboratories around the world. However, with the huge surge in diagnostic testing, laboratories began competing for the same test components and certain reagents such as RNA extraction kits became difficult to source. Consequently, to ensure a robust, resilient diagnostic service with an increased capacity, Hampshire Hospitals NHS Foundation Trust (HHFT) sought to diversify the portfolio of testing strategies by exploring alternative chemistries which have separate reagent supplier pathways to those of rRT-PCR, and which also permit direct testing without the need for RNA extraction.

Reverse-transcription loop-mediated isothermal amplification (RT-LAMP) satisfied these requirements by combining reverse-transcription and autocycling, isothermal, strand displacement DNA amplification to produce a highly sensitive, versatile and robust test^5–7^. LAMP chemistry is more resistant to inhibitors than rRT-PCR, enabling simplification and even removal of extraction procedures^8^. LAMP has been applied for the detection of a wide range of pathogens, including positive-sense RNA viruses and has been used extensively in the veterinary and plant industry ^9–11^ and more recently in human diagnostics ^12–16^. Herein we describe the validation of a novel SARS-CoV-2 RT-LAMP assay which can be performed on extracted RNA, or directly from viral transport medium (VTM) taken from combined oropharyngeal and nasopharyngeal swabs (ONSwab).

## 2. Methods

### 2.1. Virus isolates and clinical specimens

Diagnostic sensitivity (DSe) and specificity (DSp) were determined using adult inpatient ONSwabs submitted to the Microbiology department at HHFT during March and April 2020, which had previously been confirmed either SARS-CoV-2 RNA positive or negative by SoC rRT-PCR. All ONSwabs were collected in Sigma Virocult^®^ medium (Medical Wire & Equipment, Corsham, UK).

Analytical sensitivity (ASe) of RNA-RT-LAMP was determined using a ten-fold dilution series of SARS-CoV-2 RNA purified from virus infected tissue culture fluid (BetaCoV/England/02/2020) obtained from Public Health England (Lot 07.02.2020) and a titration of a synthetic DNA fragment containing the SARS-CoV-2 RT-LAMP target in nuclease free water (NFW) (Integrated DNA Technologies, Coralville, United States).

ASe of Direct RT-LAMP was determined using a two-fold dilution series (1:8 to 1:2048) of VTM taken from a SARS-CoV-2 positive ONswab sample. A standard curve (Qnostics, Scotland, UK) was run on the rRT-PCR, allowing quantification of RNA in digital copies (Log_10_ dC/ml). Analytical specificity (ASp) was determined using the NATtrol™ Respiratory Verification Panel 2 (ZeptoMetrix Corporation, New York, United States) containing pathogens causing indistinguishable clinical signs to COVID-19 (n=22) and a pool of meningitis encephalitis causative agents (n=7) (Table 1).

**Table 1:** The panel of respiratory and meningitis/encephalitis pathogens used for analytical specificity.

| **Respiratory Pathogen** |
| --- |
| Coronavirus OC43 |
| Adenovirus 31 |
| Parainfluenza 4 |
| Influenza B |
| Influenza AH3 |
| Parainfluenza 3 |
| Rhinovirus 1A |
| Coronavirus 229E |
| Parainfluenza 2 |
| Adenovirus 1 |
| Coronavirus NL63 |
| Respiratory syncytial virus A2 |
| Influenza A H1N1 |
| Influenza A pandemic H1N1 |
| Parainfluenza 1 |
| *Mycoplasma pneumoniae* |
| Adenovirus 3 |
| *Bordetella pertussis* |
| *Chlamydia pneumoniae* |
| *Bordetella parapertussis* |
| Coronavirus HKUI |
| Human metapneumovirus 8 |
| **Meningitis/Encephalitis Pathogen** |
| *Neisseria meningitidis* |
| *Streptococcus agalactiae* |
| *Haemophilus influenzae* |
| Herpes simplex virus 2 |
| *Listeria monocytogenes* |
| *Parechovirus type 3* |
| Varicella zoster virus |

Repeatability, inter-operator and inter-platform reproducibility were determined using combined ONSwabs submitted to HHFT, previously confirmed as SARS-CoV-2 positive, and a SARS-CoV-2 Medium Q Control 01 positive control (Qnostics, Scotland, UK) (diluted 1 in 10 and 1 in 100).

Preliminary evaluation of Direct RT-LAMP for detection of SARS-CoV-2 in other clinical samples was performed using fourteen saliva samples collected from HHFT in-patients during May 2020 confirmed from paired ONSwabs as positive and negative for SARS-CoV-2. Collection of saliva involved the patient providing a fresh saliva sample into a 10 ml universal container. Prior to analysis the saliva was diluted 1:5, 1:10 and 1:20 in NFW.

### 2.2. RNA extraction

RNA was extracted using the Maxwell^®^ RSC Viral Total Nucleic Acid Purification Kit (Promega UK Ltd., Southampton, UK) according to manufacturer’s instructions. Briefly, a Class 1 microbiological safety cabinet (MSC) within a containment level 3 laboratory, 200 µl of sample was added to 223 µl of prepared lysis solution (including 5 µl per reaction of Genesig^®^ Easy RNA Internal extraction control, Primerdesign Ltd, Chandler’s Ford, UK). Samples were then inactivated for 10 minutes at room temperature within the MSC and 10 minutes at 56°C on a heat block before automated RNA extraction using a Maxwell^®^ RSC 48 Instrument (Promega UK Ltd., Southampton, UK). RNA was eluted in 50 µl of NFW. In the case of saliva, RNA was extracted from 200 µl of saliva diluted 1:20, as saliva volume was insufficient unless a dilution was performed.

### 2.3. Real-time reverse-transcription PCR (rRT-PCR)

rRT-PCR assays were performed in single replicates using 5 µl of RNA template with the COVID-19 genesig^®^ Real-Time PCR assay (Primerdesign Ltd, Chandler’s Ford, UK) according to the manufacturer’s guidelines, on a MIC qPCR Cycler (Bio Molecular Systems, London, UK). Single replicates were performed to ensure an adequate supply of reagents. The cycling conditions were adjusted to the following: a reverse-transcription (RT) step of 10 minutes at 55°C, a hot-start step of 2 minutes at 95°C, and then 45 cycles of 95°C for 10 seconds and 60°C for 30 seconds. The Genesig^®^ COVID-19 positive control included in the kit, a negative extraction control, and a no template control were also included on each rRT-PCR run.

### 2.4. Reverse-transcription loop-mediated isothermal amplification (RT-LAMP)

RT-LAMP reactions were performed using OptiGene Ltd. (Horsham, UK) COVID-19_RT-LAMP kits which target the *ORF1ab* region of the SARS-CoV-2 genome: (i) COVID-19_RNA RT-LAMP KIT-500 kit (for use on extracted RNA) and (ii) COVID-19_Direct RT-LAMP KIT-500 kit (for use on diluted combined ONSwabs). The COVID-19_Direct RT-LAMP KIT-500 kit contains an additional proprietary enhancing enzyme.

Each RT-LAMP reaction consisted of: 17.5 μl of RT-LAMP Isothermal Mastermix (containing 8 units of GspSSD2.0 DNA Polymerase, 7.5 units of Opti-RT reverse transcriptase and a proprietary fluorescent dsDNA intercalating dye), 2.5 μl of 10X COVID-19 Primer Mix, and 5 μl of RNA/sample. RT-LAMP reactions were performed in duplicate at 65°C for 20 mins on a Genie^®^ HT or portable Genie^®^ III (OptiGene Ltd., Horsham, UK). An exponential increase in fluorescence (ΔF) indicated a positive reaction, which was quantified by a time to positivity (Tp) value, called at the point where the fluorescence level on the amplification curve crosses the threshold of 5000. To confirm the specificity of the amplification reaction, an anneal curve was performed: RT-LAMP products were heated to 98°C for 1 min, then cooled to 80°C decreasing the temperature by 0.05°C/s.

Genie^®^ embedded software (OptiGene Ltd., Horsham, UK) was utilised to analyse RT-LAMP results and define thresholds for result calling. All RT-LAMP reactions were performed at least in duplicate, and a sample was considered positive when a Tp was observed in at least one replicate with amplification above 5000 fluorescence points and had an anneal temperature of between 81.50°C and 84.05°C with a derivative above 2500 F/°C.

For RNA RT-LAMP 5 μl of extracted RNA was added to the reaction and for Direct RT-LAMP 5 μl of VTM from the swab diluted 1:20 in NFW, or saliva diluted 1:5, 1:10 and 1:20 in NFW was added to the reaction.

### 2.5. Repeatability, inter-operator and inter-platform reproducibility

Repeatability and inter-operator reproducibility for the RNA RT-LAMP and Direct RT-LAMP were measured by running eight replicates of samples with three different operators. Inter-platform reproducibility was measured by running eight replicates of the samples across two Genie^®^ platforms. For RNA RT-LAMP, operators used the same RNA extraction for each sample; for Direct RT-LAMP operators used the same 1 in 20 dilution of a combined swab sample in NFW.

### 2.6. Statistical analysis

DSe, DSp, positive and negative likelihood ratios (LR) including 95% confidence intervals (CI), and the Cohen’s Kappa statistic (*κ*)^17^ were determined using contingency tables in R 3.6.1^18^. Assessment of the diagnostic performance was made under three scenarios: 1) “No CT cut off” (low-to-high viral load), 2) “C_T_ cut off ≤33” (moderate-to-high viral load) and 3) C_T_ cut off ≤25 (high viral load and significant risk of shedding).

To further explore the practical application of the RT-LAMP assay in clinical practice, we estimated a patient’s probability of being infected under different clinical scenarios where Direct RT-LAMP could be applied. Final diagnosis in these scenarios is given by linking the patient’s pre-test probability of infection (P_pre_) with the Direct RT-LAMP’s LRs to estimate the post-test probability of infection (P_post_). To estimate these pre- and post-test probabilities of infection a scenario-tree model was used^19^ which allowed estimation of risk-based probability estimates for scenarios where patients are: 1) symptomatic and have had no contact with a suspected or confirmed SARS-CoV-2 infected individual (risk contact), 2) symptomatic and have had risk contact(s), 3) asymptomatic with no risk contact(s) and 4) symptomatic and have had risk contact(s). A detailed explanation of the model and parameters used is provided as supplementary material. This model was built in Excel using the add-in software Poptools^20^ (Supplementary information).

## 3. Results

### 3.1. Analytical sensitivity

Using a synthetic DNA template titrated in NFW, the RNA-RT-LAMP and Direct-RT-LAMP assays were able to detect 1×10^1^ copies each, in one of two duplicates (detection limit between 1×10^1^ and 1×10^2^ copies/reaction) (Table 2). To compare the ASe of the RNA RT-LAMP with the rRT-PCR assay a 10-fold decimal dilution series of SARS-CoV-2 RNA extracted from a virus infected tissue culture media was used. The RT-LAMP detected to a dilution of 10^-3^, equivalent to a rRT-PCR C_T_ value of 36.0 (Table 1). In the case of RNA RT-LAMP the dilution with a corresponding rRT-PCR C_T_ ≤30 was detected in duplicate and C_T_ ≥30 and ≤39 were detected in one of the duplicates (Table 3).

**Table 2.** Analytical sensitivity (ASe) of RNA and Direct RT-LAMP using a synthetic DNA template

| *Template<br>copy number | RNA RT-LAMP<br>(Tp) |  | Direct RT-LAMP<br>(Tp) |  |
| --- | --- | --- | --- | --- |
|  | Replicate 1 | Replicate 2 | Replicate 1 | Replicate 2 |
| 1x10<sup>7</sup> | 03:32 | 03:31 | 03:46 | 03:45 |
| 1x10<sup>6</sup> | 04:14 | 04:15 | 04:29 | 04:30 |
| 1x10<sup>5</sup> | 04:49 | 04:49 | 05:03 | 05:00 |
| 1x10<sup>4</sup> | 05:31 | 05:31 | 05:40 | 05:41 |
| 1x10<sup>3</sup> | 06:30 | 06:48 | 06:58 | 07:12 |
| 1x10<sup>2</sup> | 09:37 | 08:50 | 07:36 | 10:20 |
| 1x10<sup>1</sup> | 12:53 | Negative | 10:32 | Negative |
| NTC | Negative | Negative | Negative | Negative |

Tp: Time to positivity in minutes and seconds (mm:ss); NTC: no template control. \*Copies/reaction, 25 µl.

**Table 3.** Analytical sensitivity (ASe) of RNA RT-LAMP

| Decimal 10-fold<br>dilution | rRT-PCR (C<sub>T</sub>) | RNA RT-LAMP (Tp) |  |
| --- | --- | --- | --- |
|  |  | Replicate 1 | Replicate 2 |
| Neat | 27.5 | 08:03 | 08:32 |
| 10<sup>-1</sup> | 30.0 | 10:44 | 11:45 |
| 10<sup>-2</sup> | 33.0 | 13:03 | Negative |
| 10<sup>-3</sup> | 36.0 | 14:29 | Negative |
| 10<sup>-4</sup> | 39.0 | Negative | Negative |

C<sub>T</sub>: Cycle Threshold; Tp: Time to positivity in minutes and seconds (mm:ss).

To compare the analytical sensitivity of the Direct RT-LAMP to the rRT-PCR assay a 2-fold decimal dilution series of SARS-CoV-2 positive VTM from a combined swab was used. The Direct RT-LAMP detected dilutions spanning 1:8 to 1:512, equivalent to a rRT-PCR C_T_ value of 22.7 (Table 3). This would equate to between 5 - 6 log_10_ digital copies (dC)/ml. The rRT-PCR detected dilutions spanning 1:8 to 1:2048 (Table 4).

**Table 4.** Analytical sensitivity (Ase) of Direct RT-LAMP

| Decimal 2-fold<br>dilution | rRT-PCR (C<sub>T</sub>) | Direct RT-LAMP (Tp) |  |  |
| --- | --- | --- | --- | --- |
|  |  | Replicate 1 | Replicate 2 | Replicate 3 |
| 1:8 | 16.55 | 06:47 | 06:25 | 06:25 |
| 1:16 | 17.31 | 07:02 | 06:38 | 06:37 |
| 1:32 | 17.80 | 07:22 | 07:03 | 07:09 |
| 1:64 | 19.04 | 07:41 | 07:53 | 07:38 |
| 1:128 | 20.03 | 08:26 | 08:42 | 09:49 |
| 1:256 | 21.73 | 08:38 | 09:18 | 09:41 |
| 1:512 | 22.65 | 09:30 | 10:02 | Negative |
| 1:1024 | 24.15 | Negative | Negative | Negative |
| 1:2048 | 24.80 | Negative | Negative | Negative |

C<sub>T</sub>: Cycle Threshold; Tp: Time to positivity in minutes and seconds (mm:ss)

### 3.2. Performance of RNA RT-LAMP

The performance of the RT-LAMP on extracted RNA was determined using 196 individual clinical samples tested in duplicate and compared to the results of the rRT-PCR (tested in single) (Figure 1). All samples with a C_T_ ≤30 were detected within 16 minutes. The overall DSe was calculated as 97% (95% CI: 90 - 99) and the overall DSp was 99% (95 - 100) (Table 5A) (positive likelihood ratio: 103.39 [14.69 – 727.57]; negative likelihood ratio: 0.03 [0.01 – 0.10]), indicating almost perfect agreement between the two assays.

**Table 5.** Overall diagnostic sensitivity (DSe) and specificity (DSp) of the RNA RT-LAMP with all rRT-PCR C_T_ values considered (A), and with a C_T_ value cut-off of ≤33 (B) and ≤25 (C).

|  |  | **rRT-PCR (no $C_T$ cut-off, 45 cycles)** |  |  |
| --- | --- | --- | --- | --- |
| **(A)** |  | Positive | Negative | Total |
| **RNA-RT-LAMP** | Positive | 86 | 1 | 87 |
|  | Negative | 3 | 106 | 109 |
|  | Total | 89 | 107 | 196 |
|  |  | DSe = 97%, DSp = 99%, K = 0.96 |  |  |
|  |  | **rRT-PCR ($C_T$ cut-off of $\leq 33$)** |  |  |
| **(B)** |  | Positive | Negative | Total |
| **RNA-RT-LAMP** | Positive | 78 | 1 | 79 |
|  | Negative | 0 | 106 | 106 |
|  | Total | 78 | 107 | 185 |
|  |  | DSe = 100%, DSp = 99%, K = 0.99 |  |  |
|  |  | **rRT-PCR ($C_T$ cut-off of $\leq 25$)** |  |  |
| **(C)** |  | Positive | Negative | Total |
| **RNA-RT-LAMP** | Positive | 36 | 1 | 37 |
|  | Negative | 0 | 106 | 106 |
|  | Total | 36 | 107 | 143 |
|  |  | DSe = 100%, DSp = 99%, K = 0.99 |  |  |

A positive RNA RT-LAMP result is indicated by a $T_p$ of <20 minutes with the correct anneal, for at least one duplicate.

**Figure 1.**
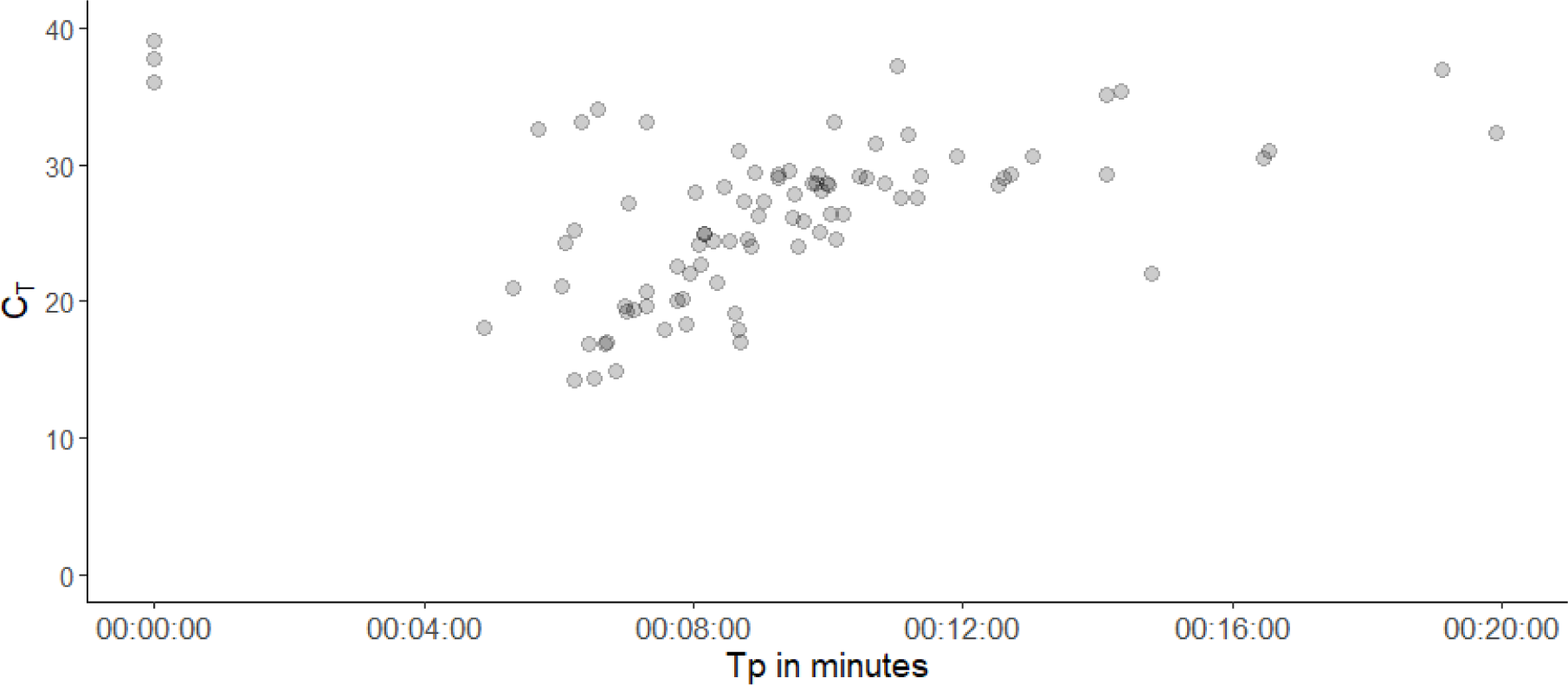
RNA RT-LAMP time to positivity (Tp: ss:mm:hh) of individual samples plotted against rRT-PCR C_T_ values. Data points represent 86 SARS-CoV-2 positive (C_T_ ≤45) clinical samples (as determined by rRT-PCR).

By employing a rRT-PCR cut-off of ≤CT 33 the RNA RT-LAMP had a DSe of 100% (95% CI: 95 - 100) and a DSp of 99% [95% CI: 95 - 100] (positive likelihood ratio: 107 [95% CI: 15.21 – 752.66]; negative likelihood ratio: 0.00 [0.00 – 0.03]), indicating almost perfect agreement between the two assays (Table 5B). By employing a rRT-PCR cut-off of ≤C_T_ 25 the RNA RT-LAMP had a DSe of 100% [95% CI: 90 - 100] and a DSp of 99% [95% CI: 95 - 100] (positive likelihood ratio: 107.00% [95% CI: 15.21 – 752.66]; negative likelihood ratio: 0.00% [0.00 – 0.05]), indicating almost perfect agreement between the two assays (Table 5C).

### 3.3. Performance of Direct-RT-LAMP

To perform RT-LAMP directly from the swab VTM a series of dilutions were evaluated comprising 1:5, 1:10, 1:20 and 1:40 in NFW. The optimal dilution whereby inhibition was limited, but Tp was maximised was determined to be 1:20 (data not shown). The DSe and DSp of the Direct-RT-LAMP assay was determined using 119 individual clinical samples diluted at 1 in 20 in NFW and compared to the results of the rRT-PCR (tested in single) (Figure 2). All samples with a C_T_ ≤30 were detected within 14 minutes.

**Figure 2.**
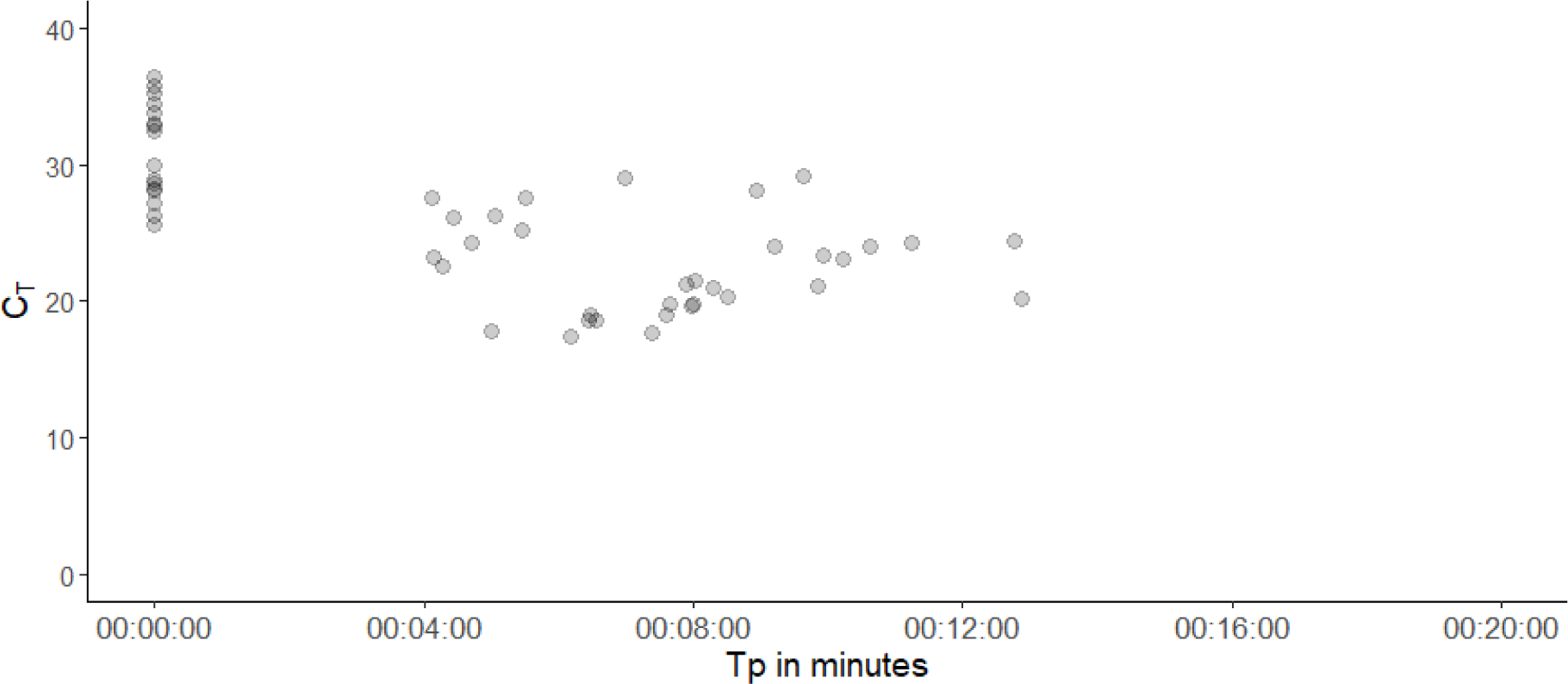
Direct RT-LAMP time to positivity (Tp: ss:mm:hh) plotted against rRT-PCR C_T_. Data points represent 49 SARS-CoV-2 positive clinical samples (as determined by rRT-rPCR).

The overall DSe of Direct-RT-LAMP was 67% [95% CI: 52 - 80] and the overall DSp was 97% [95% CI: 90 - 100], positive likelihood ratio: 23.57% [95 CI: 5.93 – 93.68], negative likelihood ratio: 0.34 [95% CI: 0.22 – 0.50], with substantial agreement between the two assays (Table 6A).

**Table 6.**
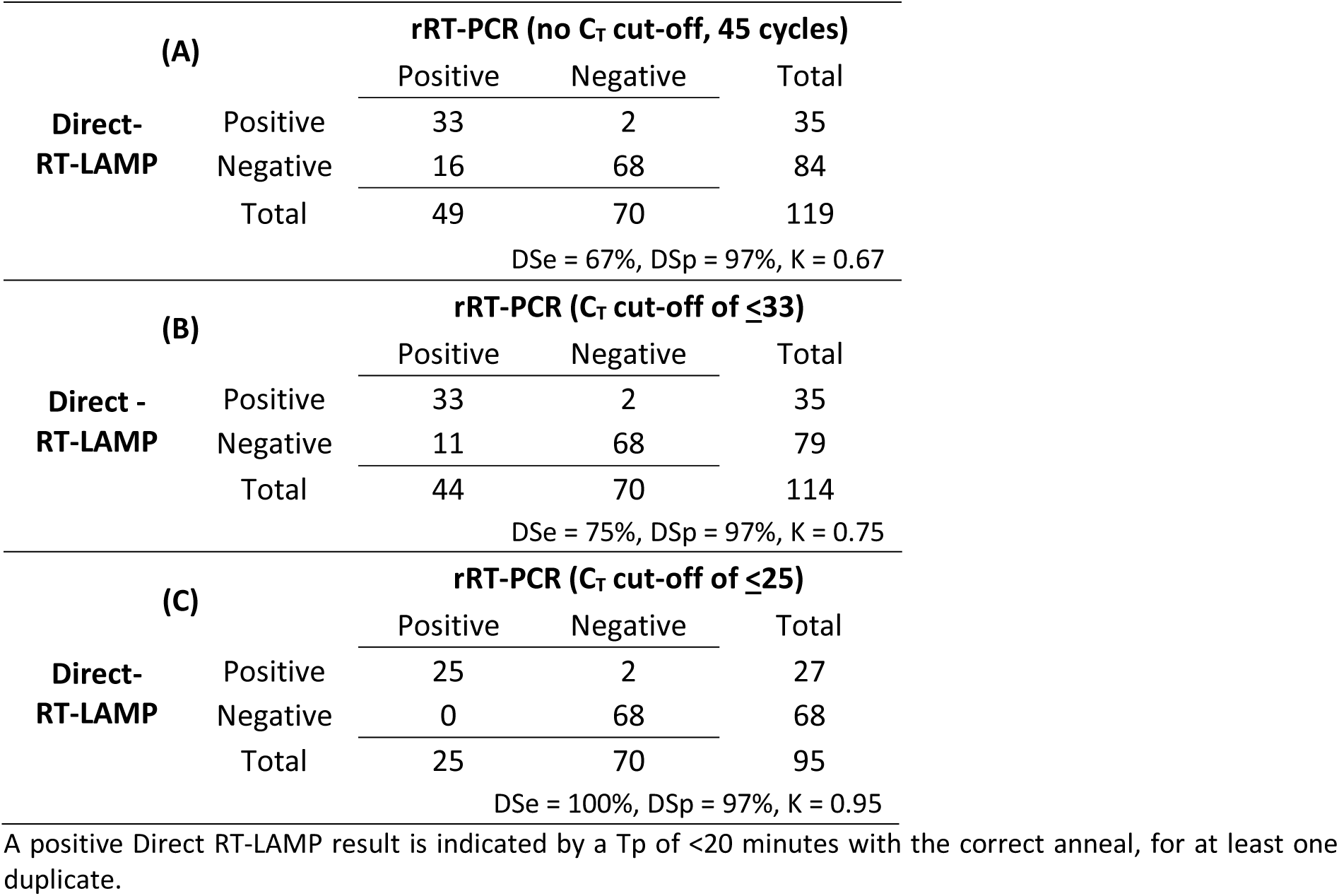
Overall diagnostic sensitivity (DSe) and specificity (DSp) of the Direct-RT-LAMP with all rRT-PCR C_T_ values considered (A), and with a C_T_ value cut-off of ≤33 (B) and ≤25 (C).

|  |  | **rRT-PCR (no $C_T$ cut-off, 45 cycles)** |  |  |
| --- | --- | --- | --- | --- |
| **(A)** |  | Positive | Negative | Total |
| **Direct-RT-LAMP** | Positive | 33 | 2 | 35 |
|  | Negative | 16 | 68 | 84 |
|  | Total | 49 | 70 | 119 |
|  |  | DSe = 67%, DSp = 97%, K = 0.67 |  |  |
|  |  | **rRT-PCR ($C_T$ cut-off of $\leq 33$)** |  |  |
| **(B)** |  | Positive | Negative | Total |
| **Direct - RT-LAMP** | Positive | 33 | 2 | 35 |
|  | Negative | 11 | 68 | 79 |
|  | Total | 44 | 70 | 114 |
|  |  | DSe = 75%, DSp = 97%, K = 0.75 |  |  |
|  |  | **rRT-PCR ($C_T$ cut-off of $\leq 25$)** |  |  |
| **(C)** |  | Positive | Negative | Total |
| **Direct-RT-LAMP** | Positive | 25 | 2 | 27 |
|  | Negative | 0 | 68 | 68 |
|  | Total | 25 | 70 | 95 |
|  |  | DSe = 100%, DSp = 97%, K = 0.95 |  |  |

A positive Direct RT-LAMP result is indicated by a $T_p$ of <20 minutes with the correct anneal, for at least one duplicate.

The DSe when a rRT-PCR C_T_ value cut-off of ≤33 or ≤25 was utilized, increased to 75% [95% CI: 60 - 87] and 100% [95% CI: 86 - 100] respectively (Table 6B and 6C). Positive likelihood ratios were 26.25 [95% CI: 6.63 – 103.98] and 35 [95% CI: 8.93 – 137.18], respectively, and negative likelihood ratio were 0.26 [95% CI: 0.15 – 0.43] and 0.00 [95% CI: 0.00 – 0.08] respectively. There was substantial agreement using a C_T_ cut off ≤33 and almost perfect agreement using a C_T_ cut off ≤25. When the ASe was determined independently from the DSe using a dilution series of SARS-CoV-2 patient swab VTM, it was noted that a C_T_ value of 24.2 and 24.8 were not detected by Direct RT-LAMP. This contrasts with the results from the DSe evaluation when these range of C_T_ were detected. A C_T_ of 24 directly from VTM is not necessarily comparable to a C_T_ of 24 derived from a serially diluted swab sample and this likely reflects the difference observed. Using a standard curve to measure genome copies was not performed for DSe, but it was used for ASe.

The incorporation of subsequent confirmatory rRT-PCR testing to verify a negative Direct RT-LAMP result increased the overall DSe of this pipeline to 99%, with a DSp of 98.4%. ASp was determined using a panel of respiratory pathogens, including four seasonal coronaviruses Direct-RT-LAMP. No cross reactivity was observed.

A selection of paired ONSwab and saliva samples were compared to evaluate saliva as a potential diagnostic matrix for SARS-CoV-2 detection (Table 7). The ONSwab samples ranged in C_T_ value from 18.6 to 35.8 when the rRT-PCR was performed on neat VTM and ranged in Tp from 06:09-11:36 minutes. Direct RT-LAMP detected SARS-CoV-2 in all samples (n=4) with a C_T_ ≤25. Direct RT-LAMP did not detect SARS-CoV-2 in ONSwab VTM with a C_T_ ≥25 (n=4). SARS-CoV-2 was detected in only two of the paired saliva swabs in all dilutions (1:5, 1:10, 1:20) for one sample and in two dilutions (1:5 and 1:10) for the other sample. All four rRT-PCR negative samples were negative by Direct RT-LAMP both in the ONSwabs and in the saliva samples.

**Table 7.** Comparison between swab and saliva detection of SARS-CoV-2 using Direct RT-LAMP

| Sample | C<sub>T</sub> from neat ONSwab | Tp 1:20 ONSwab | C<sub>T</sub> from 1:20 Saliva | Tp 1:5 Saliva | Tp 1:10 Saliva | Tp 1:20 Saliva |
| --- | --- | --- | --- | --- | --- | --- |
| 1 | 35.81 | Negative | Negative | Negative | Negative | Negative |
|  |  | Negative |  | Negative | Negative | Negative |
| 2 | 17.44 | 06:14 | 30.36 | Negative | Negative | Negative |
|  |  | 06:09 |  | 11:48 | 10:55 | Negative |
| 3 | 28.97 | Negative | 31.94 | Negative | Negative | Negative |
|  |  | Negative |  | Negative | Negative | Negative |
| 4 | 34.46 | Negative | 31.04 | Negative | Negative | Negative |
|  |  | Negative |  | Negative | Negative | Negative |
| 5 | 24.26 | 10:52 | 24.91 | Negative | Negative | Negative |
|  |  | 11:36 |  | Negative | Negative | Negative |
| 6 | 18.97 | 06:31 | 25.17 | 07:48 | 08:07 | 07:30 |
|  |  | 06:26 |  | 08:00 | 09:18 | 08:53 |
| 7 | 18.56 | 06:23 | 31.74 | Negative | Negative | Negative |
|  |  | 06:32 |  | Negative | Negative | Negative |
| 8 | 32.46 | Negative | Negative | Negative | Negative | Negative |
|  |  | Negative |  | Negative | Negative | Negative |
| 9 | Negative | Negative | Negative | Negative | Negative | Negative |
|  |  | Negative |  | Negative | Negative | Negative |
| 10 | Negative | Negative | Negative | Negative | Negative | Negative |
|  |  | Negative |  | Negative | Negative | Negative |
| 11 | Negative | Negative | Negative | Negative | Negative | Negative |
|  |  | Negative |  | Negative | Negative | Negative |
| 12 | Negative | Negative | Negative | Negative | Negative | Negative |
|  |  | Negative |  | Negative | Negative | Negative |
| 13 | Negative | Negative | Negative | Negative | Negative | Negative |
|  |  | Negative |  | Negative | Negative | Negative |
| 14 | Negative | Negative | Negative | Negative | Negative | Negative |
|  |  | Negative |  | Negative | Negative | Negative |

C<sub>T</sub>: Cycle Threshold; Tp: Time to positivity in mm:ss; ONSwab: Combined oro and nasopharyngeal swab

### 3.4. Repeatability, inter-operator and inter-platform reproducibility

When it comes to repeatability and inter-operator reproducibility, 100% of the replicates were detected for each sample by the three operators. The percentage coefficient of variation (%CV) was below 10 both when comparing within and between operators (Table 8). When comparing between platforms, 100% of the replicates were detected on both the Genie^®^ HT and Genie^®^ III, with the %CV below 10 (Table 9).

**Table 8.** Repeatability and inter-operator reproducibility

| Sample | Reaction | rRT-PCR<br>(C<sub>T</sub>) | Mean Tp<br>(% coefficient of variation) |  |  | Reproducibility<br>between<br>operators |
| --- | --- | --- | --- | --- | --- | --- |
|  |  |  | Operator 1 | Operator 2 | Operator 3 |  |
| ONSwab<br>(RNA diluted 1/10) | RNA-RT-LAMP | 21.64 | 05:16 (0.61) | 05:03 (0.51) | 04:48 (0.26) | 05:02 (4.72) |
| Qnostics positive control<br>(diluted 1/10) | RNA-RT-LAMP | 24.94 | 06:05 (1.44) | 05:54 (1.22) | 05:41 (1.08) | 05:54 (3.38) |
| Qnostics positive control<br>(diluted 1/100) | RNA-RT-LAMP | 29.27 | 07:08 (4.24) | 07:09 (2.72) | 06:48 (6.06) | 07:02 (2.85) |
| ONSwab<br>(Swab VTM diluted 1/20) | Direct-RT-LAMP | 25.50 | 07:11 (7.28) | 07:45 (8.48) | 07:38 (7.40) | 07:31 (3.94) |

VTM: Viral Transport Medium. C<sub>T</sub>: Cycle Threshold; ONSwab: Combined oro and nasopharyngeal swab; Tp: Time to positivity in mm:ss

**Table 9.** Inter-platform reproducibility

| Sample | Reaction | rRT-PCR<br>(C<sub>T</sub>) | Mean Tp<br>(% coefficient of variation) |  | Reproducibility<br>between platforms |
| --- | --- | --- | --- | --- | --- |
|  |  |  | Genie® HT | Genie® III |  |
| ONSwab<br>(RNA diluted 1/10) | RNA-RT-LAMP | 21.64 | 05:16 (0.61) | 04:49 (0.61) | 05:02 (6.37) |
| Qnostics positive control<br>(diluted 1/10) | RNA-RT-LAMP | 24.94 | 06:05 (1.44) | 05:39 (1.33) | 05:52 (5.35) |
| Qnostics positive control<br>(diluted 1/100) | RNA-RT-LAMP | 29.27 | 07:08 (4.24) | 06:46 (4.54) | 06:57 (3.71) |
| ONSwab<br>(Swab VTM diluted 1/20) | Direct-RT-LAMP | 25.50 | 07:11 (7.28) | 06:41 (4.40) | 06:56 (5.16) |

VTM: Viral Transport Medium. C<sub>T</sub>: Cycle Threshold; ONSwab: Combined oro and nasopharyngeal swab; Tp: Time to positivity in mm:ss

### 3.6 Linking pre- and post-test probability of infection

The practical application of using Direct RT-LAMP during the growing phase of an epidemic where the prevalence of infection is around 0.14 (14%) (Supplementary Information 1) was modelled. In practice a clinical team will assess patients who have clinical signs (symptomatic) or not (asymptomatic) and those that have either had contact or not with sick or infected individuals (risk contact). These patients all have different risks and therefore different pre-test probabilities of being infected (Figure 3). Pre- and post-test probabilities of infection are presented for different risk categories of patients and different risk categories of viral shedding levels (no C_T_ cut off, C_T_ ≤33, C_T_ ≤25) (Figure 3). For example, consider a symptomatic patient who had no risk contact. As shown in Figure 3, the pre-test probability that they are infected is on average 0.19 (19%), after testing positive in the Direct RT-LAMP test, the (post-test) probability of this patient being infected increased to 0.81 (81%). On the other hand, if the Direct RT-LAMP result was negative the probability of the patient being infected decreases to 0.07 (7%). Assuming this probability is considered too high, the clinical team would recommend isolation until confirmatory diagnosis is obtained.

**Figure 3.**
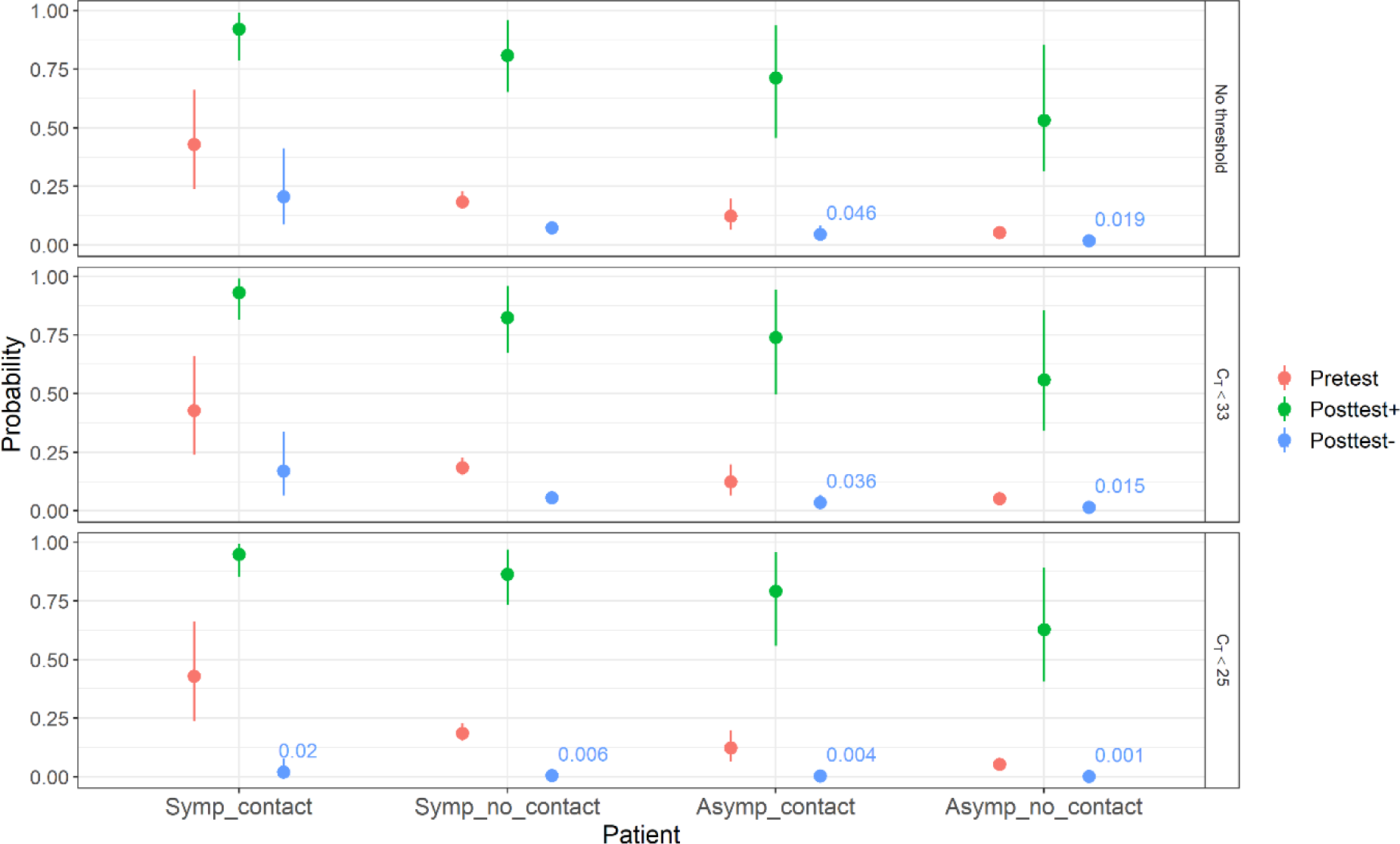
Pre- and post-test probability of infection and the use of Direct RT-LAMP. Probabilities are shown as mean (points) and 95% confidence intervals (error bars). Four risk categories of patients are considered (x axis): 1) Symp_contact = symptomatic patient with history of contact with an infected person, 2) Symp_no_contact = symptomatic patient who had no contact with an infected or sick person, 3) Asymp_contact = asymptomatic patient with history of contact with an infected Aldermaston Roadperson and 4) Asymp_no_contact = asymptomatic patient who had no contact with an infected or sick person. Post-test probability negative values ≤ 0.05 are also shown in the figure.

Consider now an asymptomatic patient with a confirmed contact awaiting a test result. The pre-test probability of this patient is 0.12 (12%), after a negative Direct RT-LAMP result the post-test probability of this patient being infected is 0.05 (5%). The clinical team, before sending the sample for confirmatory testing, may look at the post-test probability of this patient shedding moderate to high levels of virus if they were infected (C_T_ ≤ 33, C_T_ ≤25). These probabilities are lower than 0.05 (5%) (Figure 3) so the clinical team may consider these probabilities low and infer that the patient does not represent a risk for spreading infection, and diagnose the patient as “not infected”. These kinds of decisions may be necessary when there are limited diagnostic resources available.

## 4. Discussion

This study describes the development and validation of a rapid, accurate and versatile SARS-CoV-2 RT-LAMP assay. This assay demonstrates excellent concordance with rRT-PCR when performed on extracted RNA and when used directly on diluted VTM can detect samples with a high viral load which would be considered significant for viral transmission^21^. No cross reactivity was observed against common respiratory pathogens including seasonal coronaviruses.

The overall DSe of the RNA RT-LAMP assay was calculated as 97% and the overall DSp was 99% with all samples of C_T_ ≤30 detected within 16 minutes. We therefore recommend that when using RNA RT-LAMP, the length of the assay should be a maximum of 16 minutes to avoid detection of degraded nucleic acid which may be derived from the clinical sample or the environment^22^.

A shortage in the supply of RNA extraction reagents was a critical rate-limiting step affecting COVID-19 diagnostic capacity, thus the ability to bypass this step and test directly from swab has significant advantages. Various simple sample preparation methods have been reported which can circumvent RNA extraction, including the use of syringe filtration, Chelex™ 100, dilution in NFW, or a heat step, among others^23–26^. In this study the best performance for Direct RT-LAMP was achieved using a 1:20 dilution of VTM in NFW. This sample preparation method is simple and quick to perform (<5 mins) and does not require any additional equipment, therefore it is well-suited for near-patient testing.

Recent publications have demonstrated that there is a strong correlation between rRT-PCR C_T_ values and the ability to recover live virus, and therefore it is unlikely that patients providing samples with high C_T_ values pose a high risk of transmission^21^. One previous study demonstrated that live virus could only be recovered reliably from samples with a C_T_ between 13 to 17, when using a rRT-PCR targeting the *E* gene^21^. Additionally, the ability to recover live virus then dropped progressively with virus unrecoverable from samples with a C_T_ above 33^21^. Bullard and colleagues^27^ found no virus was recoverable from clinical samples taken from symptomatic patients with rRT-PCR (targeting the *E*-gene) C_T_ values of >24. In the same study^27^ each unit increase in C_T_ value corresponded to a 32% decrease in the odds of recoverable live virus. Consequently, as the risk of SARS-CoV-2 transmission is still not fully understood, a range of C_T_ cut-off values were set in our study, to understand in particular the performance of the Direct-RT-LAMP assay at different viral loads. The overall DSe of Direct RT-

LAMP was 67%, however, when setting C_T_ cut-offs of ≤33 (low-medium viral load) and ≤25 (high viral load and significant risk of shedding) the Direct RT-LAMP DSe increased to 75% and 100%, respectively. DSp was unchanged and remained at 97%. As no samples were detected beyond 14 minutes, we recommend that when using Direct RT-LAMP the length of the assay should be a maximum of 14 minutes to avoid detection of degraded nucleic acid which may be derived from the clinical sample or environment^22^.

The ability to detect patients with high viral load (C_T_ ≤25) directly from diluted swabs, demonstrates significant potential for the use of Direct RT-LAMP for the rapid diagnosis of symptomatic patients and also for rapid screening of asymptomatic individuals. This is largely supported by studies reporting similar viral loads in asymptomatic and symptomatic patient groups^28–31^, albeit not universally^32,33^. As with any diagnostic test, when it comes to the clinical application of Direct RT-LAMP, the pre-test probability of infection, based on clinical context and disease prevalence in the test subject or population, must be considered together with limitations of assay performance. We have provided a model, utilising published data on disease transmission from elsewhere, to illustrate the impact of pre-test probability on the positive predictive value (PPV) and negative predictive value (NPV) of Direct RT-LAMP in different scenarios. Depending on factors such as assay function (diagnosis vs screening), disease prevalence, patient group, setting and available resources, and their impact on PPV and NPV, further confirmation by a negative verification step may be considered desirable. It should be noted that the estimates of pre- and post-test probabilities of infection in this study were made only as an example of, and to help understand the use of the Direct RT-LAMP in practice. These estimates were based on crude approximations of the model’s parameter values (Supplementary information) and we encourage the readers who would like to use this model, to adjust the model and use parameter values that best suits the epidemiological situation of the country/region where the test would be applied.

Rapid testing of symptomatic SARS-CoV-2 positive patients within healthcare facilities allows their rapid isolation or cohorting, significantly reducing onward transmission and improving bed management and patient flow. Additionally, screening of asymptomatic patient groups or at the community level may enable the rapid identification of those with high viral loads who may pose a high risk of onward transmission. This would allow for swift public health intervention with instruction to self-isolate/quarantine and rapid track and trace methods deployed - essential in surveillance programmes aiming to reduce the reproductive number (R_0_) and spread of the disease in a community.

Direct RT-LAMP offers speed, robustness and portability making it attractive as an option for near-patient testing outside the conventional clinical laboratory, subject to the necessary risk-assessments to ensure safety of the operator^34^. Within HHFT we are exploring its application in settings such as: a multi-disciplinary non-specialist laboratory; the emergency department; primary care and care home settings. However, it must be highlighted that a test with this demonstrated sensitivity will require negative verification from a suitability sensitive diagnostic test in line with the WHO diagnostic test Target Product Profile (TPP).

In this study, clinical validation of the RT-LAMP assay took place in March, April and May 2020, largely during a period of high local COVID-19 prevalence (∼40% positivity of submitted samples) and on a limited number of samples available at the time from largely symptomatic adult patients and hospital staff. It is possible that RT-LAMP assay performance on samples from asymptomatic subjects may vary dependent on the level of detectable RNA (as a surrogate of live viral shedding) in this different patient group. Consequently, further evaluation using a larger sample set and from different scenarios is recommended, for example during periods of low prevalence or in asymptomatic individuals. Indeed, during the period of evaluation of the RT-LAMP assays, the laboratory was quickly evolving based on testing and staffing requirements, and therefore contamination of equipment or reagents occurred infrequently, which may have been the reason for the less than perfect diagnostic specificity of the RT-LAMP assays. Further evaluation within established laboratories may therefore result in an increase in diagnostic specificity, where contamination may be less likely.

Additionally, the RT-LAMP assay was validated using ONSwabs in VTM. Assay performance on a limited number of salivary samples was also explored. This preliminary analysis suggests that further research needs to be undertaken to explore saliva as a matrix for detection of SARS-CoV-2 both by rRT-PCR and Direct RT-LAMP. The drop in performance that we observed when compared to ONSwabs could be due to a number of factors causing either degradation of the RNA within the sample (e.g. salivary enzymes), or inhibition due to the complex nature of this matrix. Assay performance was not evaluated on lower respiratory tract samples or non-respiratory tract samples, and therefore future research may aim to determine the performance of both the RNA- and Direct-RT-LAMP assays using these various sample types.

The standard of care assay used as the reference standard for both RT-LAMP assays targeted the *RdRp* gene. There were two reasons for choosing the assay; one it was the assay currently validated, available and in clinical use and secondly the *RdRp* gene sits within the Orf1b region that the Optigene Ltd. assay targets making it appropriate at the time. There has been some discussion about the sensitivity of the *RdRp* gene for SARS-CoV-2 due to the poor performance of some of the early assays containing this target^35^. However, Primer Design have demonstrated *in silico* versus the submitted SARS-CoV-2 sequences throughout the pandemic that the region within *RdRp* gene that is targeted by the genesig assay is very robust as a diagnostic^36^. However, the authors would recommend that further benchmarking should be carried out versus other more sensitive Orf1ab/*RdRp* assays with additional gene targets^37^ on RT-LAMP naïve clinical sites to generate more performance data and help determine if this level of sensitivity is clinically relevant.

In our experience, during the diagnostic response to this current pandemic caused by a novel emergent pathogen (SARS-CoV-2), diversity in diagnostic platforms and routes to deliver a result based on the ability and agility to switch between methodologies has been key to allowing delivery of a resilient and sustainable diagnostic service. Factors such as: analyser availability; staff-skill mix; dynamic changes in patient groups tested or disease prevalence; and particularly in the UK; consumable and reagent supply, have highlighted the need for diagnostic services to have adaptability and capability to explore novel and alternative techniques.

## Data Availability

All data is available as required at the end of the document.

## Ethical approval

No ethical approval was required for this service evaluation study.

## 5. Acknowledgements

We would like to thank the clinical teams and Helen Denman the Microbiology laboratory manager at Hampshire Hospitals NHS Foundation Trust.

## Supplementary information

### Linking pre- and post-test probability of infection

In clinical practice diagnosis is made using a combination of the patient pre-test probability of being infected and the test result. The combination of these two will lead to an estimation of the post-test probability of infection. It is this final estimate which would help the practitioner’s decision making. In our study, the pre- and post-test probability of infection were estimated using an scenario-tree model, where different risks for infection in the estimation of the pre-test probabilities are taken into consideration^19^.

### Pre-test probability of infection

In this model the pre-test probability of infection *P*_*pre*_ was given by:

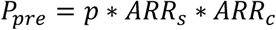

Where *p* is the prevalence of infection in the population, *ARR*_*s*_ is the adjusted risk ratio for infection of a symptomatic or asymptomatic patient and *ARR*_*c*_ is the risk ratio of a patient being infected who did have a risk contact compared with a patient who did not have a risk contact (Table S1). The ARR were calculated as follows:

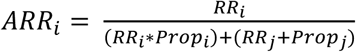

Where i, for example is an indicator for symptomatic and j is an indicator for asymptomatic. The variable *Prop* is the expected proportion (in this example) of either symptomatic or asymptomatic patients.

### Post-test probability of infection

First the pre-test odds were calculated as *Odd*_*pre*_ = *P*_*pre*_/(1 − *P*_*pre*_), then the post-test odds of infection were calculated as follows:

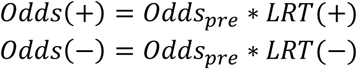

Where LRT are the positive or negative likelihood ratios of the Direct RT-LAMP:

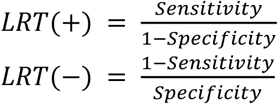

These LRT were calculated using the Direct RT-LAMP’s DSe and DSp estimates for the different viral load scenarios considered (estimated from C_T_’s: 1) “No C^T^ cut off” (high-low viral load), 2) “C_T_ cut off <33” (high-moderate viral load) and 3) C_T_ cut off <25.

Finally, the post-test odds were transformed to post-test probabilities of infection *P*_*post*_

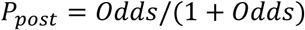

The model was implemented in Microsoft^®^ Excel^®^ using the add-in software Poptools^20^. For estimation of pre and post probabilities (mean and 95% confidence intervals), stochastic simulations of 1000 iterations were performed. Table S1 summarises the parameter values used. It should be noted that these values are crude approximations which were made only as an example of and to help understand the use of Direct RT-LAMP in practice. We encourage the readers who would like to use this model to quantify pre-and post-test probabilities of infection to better estimate the parameter values according to the epidemiological situation of the country/region where the test would be applied.

Alternatively, once the pre-test probabilities are estimated, post-test probabilities can be approximated using a Fagan nomogram^38^.

**Table S1.** Values of the parameters used for estimation of the pre- and post-test probabilities of infection

| Parameter | Value | Description | Reference |
| --- | --- | --- | --- |
| Prevalence (p) | 0.14 (14%) | Proportion of positive patients tested regardless of being symptomatic or asymptomatic | 31 |
| Proportion asymptomatic (Propasym) | Pert(0.08,0.31,0.54)a | Proportion of asymptomatic infected people.<br>Propsym = 1-Propasym | 39 |
| Risk Ratio symptomatic (RRsymp) | Pert(2.5,3.5,4.5)a | This RR was approximated as the ratio of the positive rate of symptomatic patients (0.78) to the positive rate of asymptomatic patients (0.22)<br>The reference for this RR are the asymptomatic (RRasymp = 1). | 31 |
| Risk Ratio contact risk (RRc) | Pert(1.3,2.3,3.3)a | This RR was approximated as the ratio of the positive rate when tests were done only on symptomatic patients and their contacts (0.33) to the positive rate when tests were done regardless of clinical state (0.14). The reference for this RR is the no-contact (patient had not risk contact) (RRc-no = 1). | 31 |
| Proportion risk contacts | 0.5 | For simplicity we assumed equal proportion of patients with no risk and with risk contacts |  |
| Sensitivity | Pert(0.52,0.67,0.80)a | An an example of the values of Direct RT-LAMP used directly on the sample in the scenario of "No CT cut off" (high-low viral load). Similarly Sensitivity values for the other scenarios were introduced in the model assigning a Pert distribution. | This manuscript |
| Specificity | Pert(0.90,0.97,1.00)a | See explanation given for Se. | This manuscript |

a Pert distribution (a,b,c) where a = the minimum, b = the most likely and c = the maximum values.

